# AI-Powered Radiotherapy for Resource-Limited Settings: Advancing Cervical and Prostate Cancer Treatment Planning with the Radiation Planning Assistant (RPA)

**DOI:** 10.1101/2025.10.01.25337092

**Authors:** Tucker J. Netherton, Ajay Aggarwal, Qusai Alakayleh, Beth M. Beadle, Chloe Brooks, Henriette Burger, Carlos E. Cardenas, Adrian Celaya, Sara Chacko, Christine Chung, Raphael Douglas, Daniel El Basha, Steven Frank, David Fuentes, Comron Hassanzadeh, Jonathan Helbrow, Peter Hoskin, Meena Khan, Mariana Kroiss, Alexandra Leone, Lilie Lin, Raymond Mumme, Callistus Nguyen, Quyen Nguyen, Adenike Olanrewaju, Jaganathan Parameshwaran, Julianne Pollard-Larkin, Falk Poenisch, Shalin Shah, Alan Jerel Sosa, Chad Tang, Zhiqian Yu, Lifei Zhang, Laurence E. Court

## Abstract

1.

**Purpose:** Radiotherapy treatment planning is a resource-intensive process characterized by multiple manual steps and clinical hand-offs that contribute to treatment delays and inter-observer variability. The Radiation Planning Assistant (RPA) is a web-based platform designed to deliver automated contouring and planning approaches tailored to low-resource settings. This work expands the RPA to develop and clinically validate end-to-end, AI-driven workflows for prostate and cervical cancers, designed to improve efficiency, consistency, and accessibility in low- and middle-income countries (LMICs).

**Methods:** We developed deep learning-based auto-contouring models using nnU-Net and integrated them with knowledge-based planning (KBP) models trained on curated datasets from over 1,000 prostate and 110 cervical cancer treatment plans. For prostate cancer, models were developed to accommodate prostate directed, prostate bed, and nodal treatment scenarios. Cervical cancer planning followed EMBRACE II guidelines and included pelvic and para-aortic nodal volumes. These tools were integrated into the RPA. Clinical acceptability of the auto-contours and plans was assessed retrospectively by radiation oncologists using a five-point Likert scale.

**Results:** In total, 50 test patients (40 prostate, 10 cervical) were evaluated. For prostate cancer, 70% of target auto-contours and 73% of treatment plans were clinically acceptable without edits; for cervical cancer, these rates were 80% and 80%, respectively. For prostate cancer planning, 77% of target and 98% of organ-at-risk structures met all per-protocol compliance criteria. For cervical cancer planning, all EMBRACE II protocol hard constraint criteria were met. Bowel and vaginal contours demonstrated lower performance, but these did not compromise plan quality.

**Conclusion:** We present validated, end-to-end radiotherapy planning workflows for prostate and cervical cancers that leverage the RPA’s infrastructure to streamline treatment planning in a globally accessible platform and demonstrate high clinical acceptability. By reducing reliance on specialist input, this work addresses key barriers to equitable radiotherapy access in resource-limited settings and responds to global calls from the IAEA and WHO to expand radiotherapy capacity.

**Funding:** National Institute of Health, National Science Foundation, Rising Tide Foundation, University of Texas MD Anderson Cancer Center

## 2. INTRODUCTION

Radiotherapy treatment planning is a process that involves many time-consuming steps. Contouring of the target structures on the planning CT image is typically performed by a radiation oncologist, with organs-at-risk contoured by either the radiation oncologist or a treatment planner. This is followed by treatment planning. This process, with multiple hand-offs between different members of the clinical team, can take a week or more [1]. Moreover, these manual processes are known for significant variations between users, to the extent that variations in manual contouring and planning can affect patient outcomes [2]. For these reasons, many groups have pursued the use of artificial intelligence (AI) for contouring and planning tasks, with many successful approaches reported [1,3–5].

The Radiation Planning Assistant (RPA) was designed as a web-based AI-driven tool for high-quality radiotherapy contouring and planning, with a specific interest in improving the quality of care in low- and middle-income countries. It was developed with funding from the United States National Cancer Institute Center for Global Health, as part of their Affordable Cancer Technologies (ACTs) Program [6]. The initial work was focused on cancers of the uterine cervix [7–11], post-mastectomy [12], and head and neck [13–17], with FDA clearance in 2023 and the first patient treated in South Africa in 2024 (for cervical cancer). The initial work demonstrated the interest, need, and practicality of this approach. However, it also showed opportunities for streamlining and improvement.

Although automation of individual contouring or planning tasks can give benefits in terms of efficiency, consistency, and quality, this has yet to be shown to have a significant impact on the time between CT imaging and approval of the treatment plan, or the initiation of the radiotherapy treatment. One reason for this is that much of that time is not actually active treatment planning time, but rather time waiting for the next person in the process to start their work [1]. It is important, therefore, to combine the contouring and planning tasks, such that the entire process can be completed without the need for task hand-off.

While there are some publications on true end-to-end automation in radiation therapy[12,18,19], most focus on either contouring or planning. One reason is that automation of contouring of the gross tumor volume (GTV) and high-risk clinical target volume (CTV) is a difficult task, with specifics of the volume varying widely between patients [20,21]. Significant advances have been made in the last few years, and we can expect fully automated, robust, GTV delineation to be available in the next few years. In the meantime, we propose a solution where the first task in the contouring/planning workflow is the contouring of any GTVs by the radiation oncologist, followed by all the automatable steps. These include automatic contouring of anatomic structures and treatment planning. This approach still significantly reduces hand-offs and should, therefore, have potential to significantly reduce the time between CT imaging and start of treatment.

Two treatment sites that are particularly suitable for this proposed process are prostate and cervical cancer. For both these sites, the primary targets are generally anatomical, making them good candidates for AI-based contouring. Specifically, prostate cancer targets include the prostate or prostate bed, with potential inclusion of seminal vesicles and pelvic lymph nodes; for cervical cancer, targets encompass the cervix, parametria, and vagina, with potential inclusion of the pelvic lymph nodes. In both cases, there can be boost GTV volumes in, for example, the pelvic lymph nodes. In the proposed workflow, these are the only structures that need to be manually drawn. All other steps can be automated with no user intervention. This is followed by manual review to ensure quality and prevent errors reaching the patient. Although a few researchers have reported end-to-end contouring/planning for conventional radiation therapy, including post-mastectomy breast [19], intact breast [22], palliative spine [23], and 4-field box planning for cervical cancer [10], and prostate [24,25], these approaches do not include any boost GTV regions, and in some cases testing is limited.

In this work, we build on the RPA infrastructure and both develop and integrate the tools needed for the complete end-to-end process for VMAT planning for prostate and cervical cancer patients, including AI-based contouring and planning. The tools are designed to be effective for a range of situations – for example, the patients with prostate cancer, include cases with rectal spacers, rectal balloons, and prior prostatectomy radiation. Such tools are intended increase accessibility to radiotherapy treatment planning and answer the call to the scale of up of radiotherapy resources by the IAEA, WHO, and other organizations.

## 3. MATERIALS AND METHODS

### 3.1. Prostate Auto-contouring and KBP

#### 3.1.1. Data Curation

A total of 1,069 patients who underwent intensity modulated radiotherapy for prostate carcinoma at MD Anderson Cancer Center (MDA) from 2010 to 2024 were retrospectively analyzed under a protocol approved by the MDA Institutional Review Board. Informed consents were waived due to the retrospective nature. Patients in this dataset received treatments to the prostate, prostate + nodes, prostate bed, and prostate bed + nodes. These cases also include treatment devices (e.g. rectal balloons and rectal spacers) and imaging artifacts due to metal hip implants.

#### 3.1.2. Auto-contouring models

The following contours were curated from the MDA prostate cohort: prostate, seminal vesicles (SV), prostate bed, SV-fossa, pelvic lymph node clinical target volume (CTV), rectum, sigmoid, bladder, penile bulb, femoral heads, spinal cord, cauda-equina, bowel bag, and kidneys. All curated data from the MDA cohort was evaluated for quality and adherence to ESTRO, RTOG, and NRG consensus contouring guidelines [26–29] (see Supplemental Material).

All 15 contours were placed into 5 groups for nnU-Net model training: targets, nodes, pelvic OARs, spinal OARs, and rectal spacer. The models were separated to better manage post-processing rules due to variability in structure overlap. Trained models used the 3D full resolution architecture using the standard nnU-Net settings [30]. Auto-contours for the bowel bag and kidneys utilized models previously developed for in-house clinical use [31,32].

The contour training dataset (n = 1,069) was split into training, cross-validation, and testing groups via an 80:10:10 split, respectively (Figure 1). The loss function for each model was a combination of Dice Similarity Coefficient (DSC) loss and cross-entropy loss. Training, cross validation, and testing were done on NVIDIA Tesla V100 GPUs with 32 GB VRAM. Each model was trained for 1000 epochs. Post processing rules were implemented for each specific target scenario according to institutional contouring practices and consensus guidelines (see Supplemental Material). The auto-contours were generated on 105 patients (test portion of the 80:10:10 split). The quantitative accuracy of the model was evaluated by calculating DSC and 95% Hausdorff Distance (HD95) for ground truth vs auto-contours.

**Figure 1.**
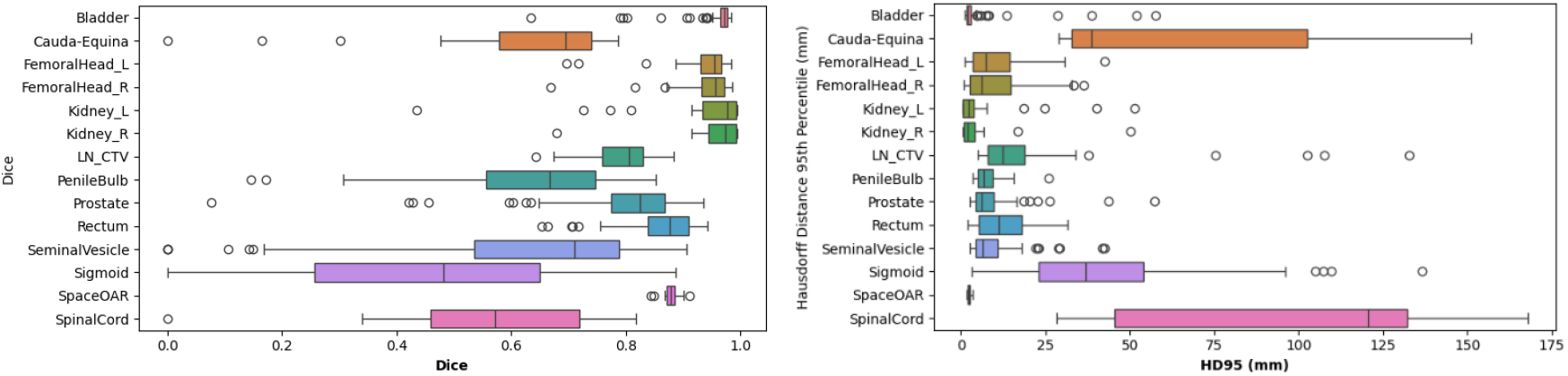
DSC (A) and 95^th^ percentile Hausdorff (B) values for prostate cancer target and OAR/other structures.

**Figure 2.**
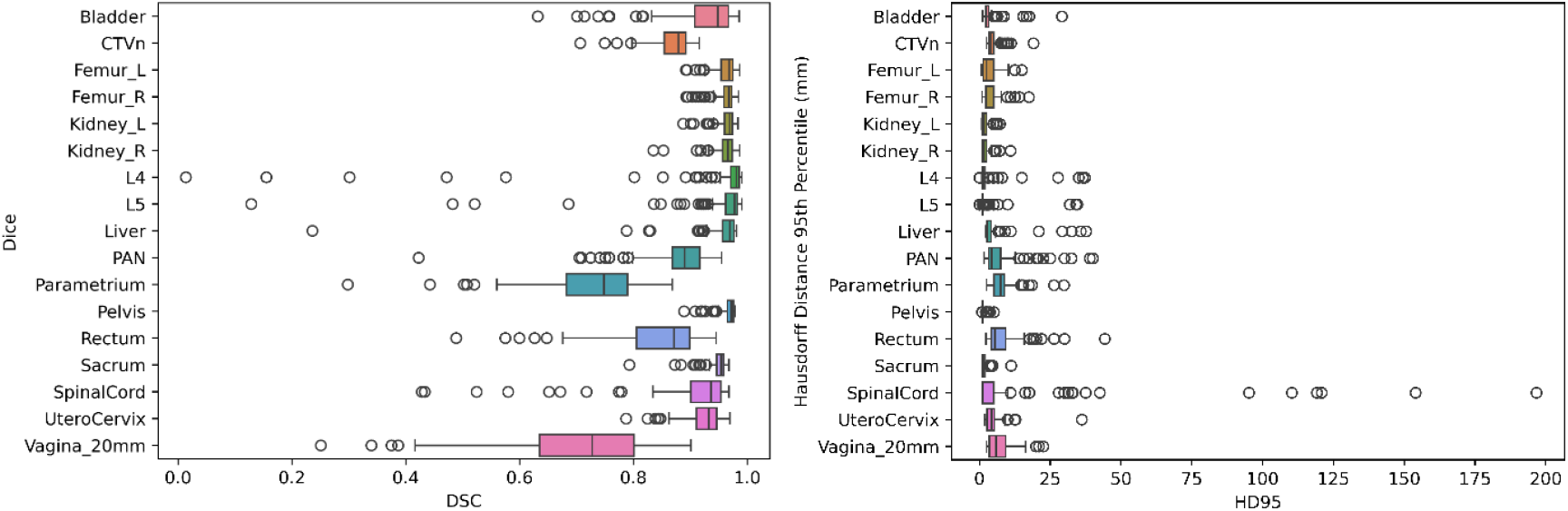
DSC (A) and 95^th^ percentile Hausdorff (B) values for cervical cancer target and OAR structures.

**Figure 3.**
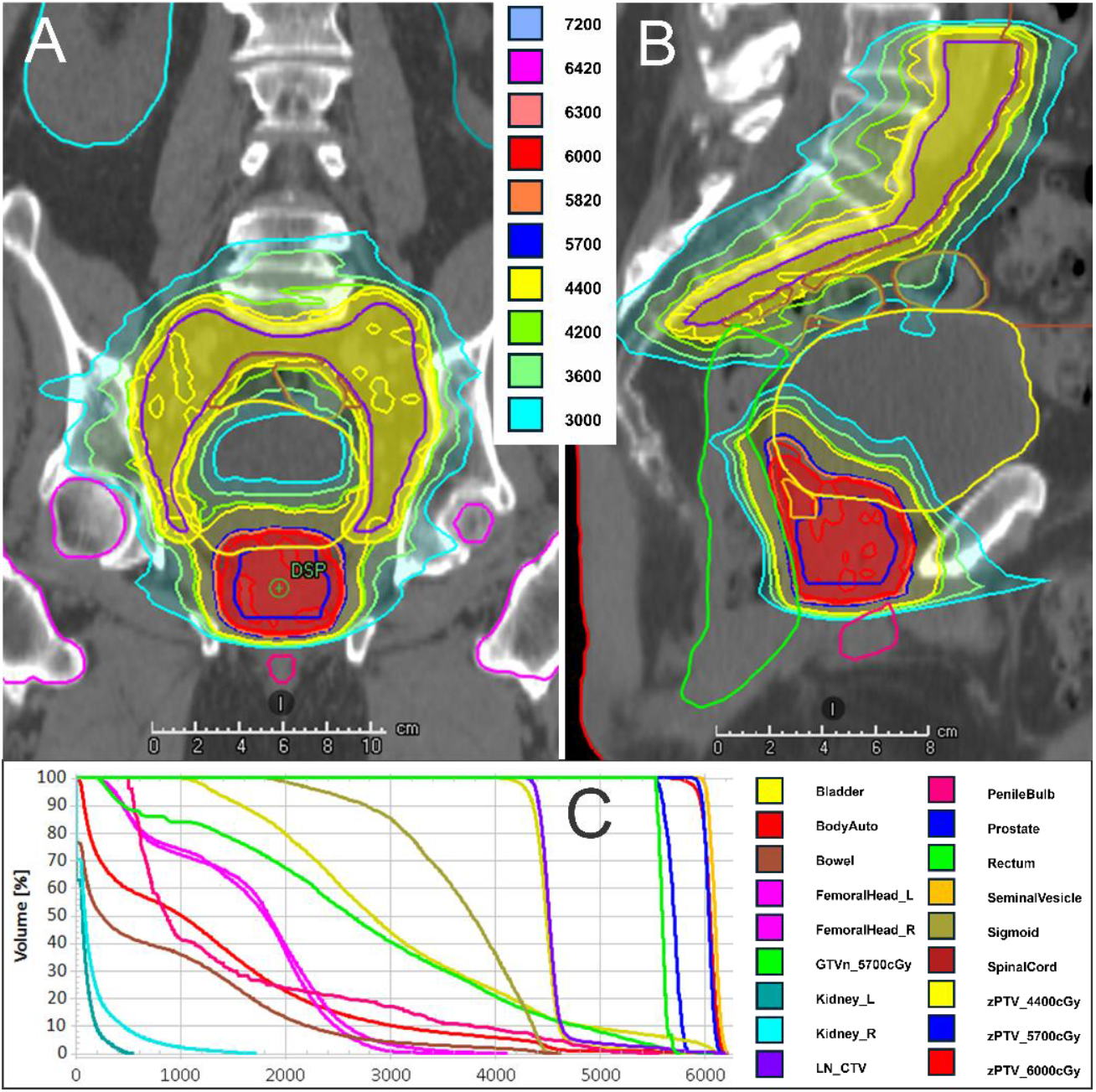
RPA prostate cancer treatment plan made using target and OAR auto-contours. This patient includes the nodal region in the PTV_4400 and includes treatment of the seminal vesicles in the PTV_6000.

**Figure 4.**
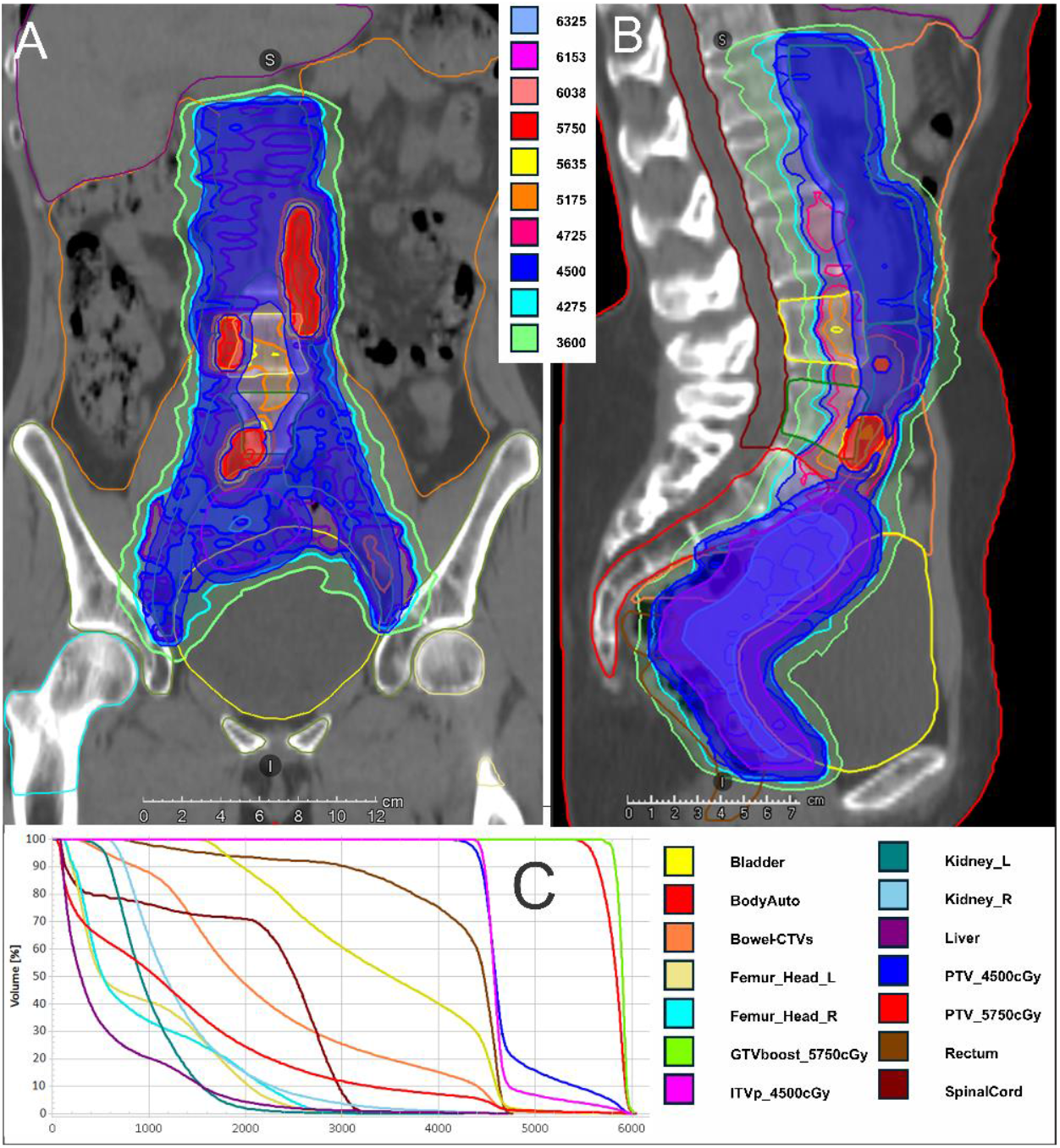
RPA cervical cancer treatment plan made using target and OAR auto-contours (manual upload of GTVBoost). This patient includes the para-aortic region in the PTV_4500.

#### 3.1.3. Knowledge-Based Planning Model

Knowledge-based planning (KBP) models were developed for prostate and prostate + nodes using RapidPlan (Varian Medical Systems, v15.6.06) in a previous work by Chung et al [15]. Briefly, plans included in each model were retrospectively selected from MDA patients that were definitively treated with radiotherapy for prostate cancer. Each case was reviewed for consistency (e.g., modality, prescription dose, extent of disease) and plan quality (e.g., OAR sparing, target coverage and conformality). The dataset excluded patients with unique setups or targets, previous treatment, metal implants or contrast within treatment field. The model structures included targets (e.g., PTV primary, PTV nodes, PTV boost), clinical OARs (e.g., bladder, bowel, femurs, penile bulb, rectum, sigmoid), and planning structures (e.g., planning risk volumes, target ring structures). The prostate model was trained on 95 cases and validated on 38 cases. The prostate + nodes model was trained on 37 cases and validated on 20 cases. These previously developed KBP models are evaluated in this work and use an end-to-end process which utilizes the auto-contours for targets and OARs from section 3.1.2.

### 3.2. Cervix Auto-contouring and KBP

#### 3.2.1. Data Curation

Our previous work in the design and validation of solutions for automated contouring and treatment planning for cervical cancer are extensive [9,10,15,19,33]. While these works have developed convolutional neural networks and knowledge-based planning approaches using training data from MD Anderson Cancer Center, the latest iteration will develop contouring and planning approaches based on EMBRACE II guidelines[34]. A total of 110 patients who underwent intensity modulated radiotherapy for gynecologic malignancies were retrospectively analyzed under a protocol approved by an Institutional Review Board. Informed consents were waived due to the retrospective nature. All patients had an intact uterus at the time of treatment planning.

To curate ground truth segmentations for auto-contouring model training, a radiologist manually contoured 17 organs-at-risk (OAR’s) and clinical target volumes (CTVs) on 100 patients following guidelines from the EMBRACE II protocol. The data (CT and gross nodal GTVs) from the ten remaining patients, previously treated from 2021-2023, were withheld for end-to-end testing of the RPA.

#### 3.2.2. Auto-contouring Models

The publicly available nnU-Net framework was used to train a full resolution, 3D segmentation model using the Dice + cross entropy loss [30]. Left and right mirror augmentations were turned off to prevent mislabeling of left and right structures (i.e. femurs and kidneys) and the model was trained for 1000 epochs. All of the training data was utilized, and 5-fold cross validation results were reported. The reported metrics were Dice Similarity Coefficient (DSC) and 95^th^ percentile Hausdorff distance.

#### 3.2.3. Knowledge-Based Planning Model

A cervical cancer specific KBP model was developed using the same methods described in Section 2.1.3. See Chung et al. 2024 for full details [15]. The model structures included targets (e.g., PTV primary, PTV nodes, PTV boost), clinical OARs (e.g., bladder, bowel, liver, femurs, kidneys, rectum, cord), and planning structures (e.g., target rings, skin fold avoid, normal tissue avoids). The model was trained on 120 cases and validated on 75 cases. In this work, this previously developed model was modified to suit the EMBRACE II planning objectives while using auto-contouring for all targets (except for contours of gross nodal disease) and OARs. A plan normalization value of 102 was applied to the PTV. Also to suit the Embrace II protocol, a mean dose limit of 15Gy to the kidneys was added. Because the end-to-end workflow uses only CT imaging, the “standardized margin” approach for generating ITV and PTV was applied per EMBRACE II recommendations.

### 3.3. Integration into the Radiation Planning Assistant

The Radiation Planning Assistant (FDA 510k cleared) was developed with funding from the NIH Center for Global Health Affordable Cancer Technologies (ACTs) program; it was created to provide no-cost automated treatment planning services for radiation clinics in LMICs [35]. The RPA requires a center to provide a patient’s computed tomography (CT) images and a prescription form from the physician; it then provides a high-quality, patient-specific set of contours and treatment plan. The RPA is a web-based system, requiring no hardware or software installation from the LMIC center, and prior work has shown that it could reduce manual effort required per patient *from days to minutes* according to a recent study by an RPA collaborating clinic in Tanzania [33].

The end-to-end workflows for prostate and cervical cancer treatment planning incorporate auto-contouring from nnU-Net and treatment planning from KBP models.

**Figure 3.**
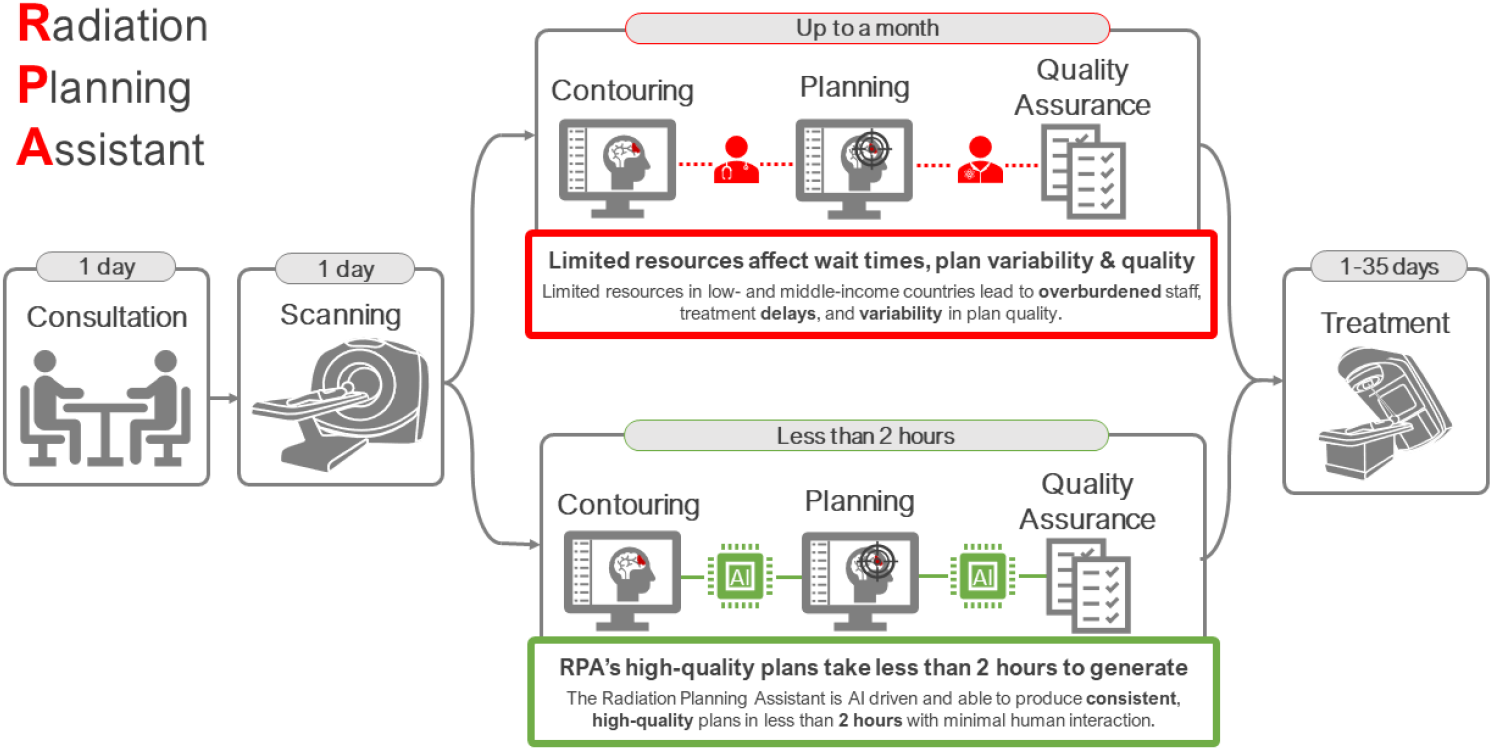
The RPA workflow. The workflow in red (top) depicts a treatment planning workflow with staff performing manual contouring, planning, and quality assurance tasks. The workflow in green (bottom) depicts a workflow where AI performs all treatment planning tasks.

### 3.4. Determination of Clinical Acceptability

Using the RPA, auto-contours and KBP plans were generated end-to-end on 50 patients (10 for cervical cancer and 40 for prostate carcinoma). These 50 were test set cases that were separate from the training sets from each auto-contouring model. For the prostate, treatment plans were made for a range of scenarios (prostate directed = 14, prostate directed + nodes = 11, prostate bed = 6, prostate bed + nodes = 9). The contours scored for clinical acceptability include prostate, seminal vesicles, rectum, sigmoid, bladder, femoral heads, penile bulb, CTV nodal, bowel bag, spinal cord, and cauda equina. Three radiation oncologists from two institutions evaluated and scored the auto-contours and plan quality on a five-point Likert scale according to Baroudi et al. (see supplementary table). Physicians 1, 2, and 3 scored 11, 5, and 24 treatment plans, respectively. In addition, dosimetric criteria for per-protocol and variation acceptable limits were evaluated for all PTVs and OARs according to NRG-GU009. It is important to note that while only 23 of the 40 plans had NRG-GU009 criteria tabulated (a limitation of this study), all plans were dosimetrically assessed and scored by a physician.

For the cervix, 5 patient plans included treatment of the para-aortic nodes (PAN) and 5 excluded the treatment of the PAN. Dose prescriptions were set according to EMBRACE II guidelines. Contours scored for clinical acceptability include nodal and primary CTV, PAN, ITV45, PTV45, rectum, bladder, bowel bag, spinal cord, femurs, kidneys, sacrum, L4, L5, and liver. The same five-point Likert scale mentioned above was used to score auto-contour and plan quality. Physicians from the Radiotherapy Trials Quality Assurance (RTTQA) group scored the cervical cancer auto-contours and plan quality. Dosimetric criteria for OARs and targets were set according to EMBRACE II guidelines. The RTTQA group is a centralized resource providing national radiotherapy QA programs for all United Kingdom trials that include a radiotherapy component.

## 4. RESULTS

### 4.1. Prostate: Auto-contouring

Target structures (prostate, seminal vesicles, and LN CTV) had average DSC ranging from 0.62 to 0.79 and HD95 values ranging from 8.28 mm to 19.6 mmm, respectively. All OARs and structures had average DSC and HD95 values ranging from 0.44 to 0.96 and 2.37 mm to 100.25 mm, respectively. For the intact, non-surgical setting, the prostate and seminal vesicles average DSC was 0.82 and 0.71 and the average HD95 were 7.60 mm and 6.64 mm, respectively. The average DSC of the prostate fossa and seminal vesicle fossa were 0.71 and 0.28 and the average HD95 were 11.0 mm and 24.1 mm, respectively. Low performing outliers from spinal cord, cauda equina, and sigmoid were from difference between the caudal and cranial extent of the contours (for ground truth versus predicted contours).

### 4.2. Cervix: Auto-contouring

Structures used to create the planning target volume (PAN, parametria, uterocervix, vagina_20mm, and CTVn) had average DSC and HD95 values ranging from 0.70 to 0.92 and 4.74 mm to 8.02 mm, respectively. All OARs had DSC and HD95 values ranging from 0.85 to 0.97 and 1.32 mm to 12.93 mm, respectively. All tabular data of quantitative results is available in the Supplementary Material. Low performing outliers from the L4 and L5 vertebra were caused due to lumbarization/sacralization. Furthermore, an outlier from the liver was present due to the liver being barely visible at the cranial portion of the scan.

### 4.3. Determination of Clinical Acceptability

For the end-to-end prostate solution, physicians 1,2, and 3 scored 11, 5, and 24 treatment plans, respectively. These treatment plans included both the auto-contours and radiotherapy plan (made on the auto-contours). Edits were not made to contours or plans, they were only scored. Overall, prostate target contouring needed at most minor edits (a score of 3) in both the prostate directed and prostate bed settings. Minor edits were those judged to be made in less time than starting from scratch or are expected to have minimal effect on treatment outcome. For prostate directed, the treatment plan quality achieved higher scores than the prostate bed scenario (Table 3). For the prostate directed and prostate directed + nodes scenario, plan quality was acceptable as is for 86% and 91% of cases, respectively. For the postoperative scenario with and without nodes, these scores were 44% and 50%, respectively. However, 98% of plans were acceptable with minor edits, regardless of the scenario (Table 3). A common remark for many of the plans was that hotspot needed to be reduced below 105%. The bowel bag was the structure that was consistently rated with lower clinically acceptable scores, and was overall, only acceptable without edits for 14% of plans. All other OAR structures received higher marks.

**Table 3.** Physician scores for contour and plan quality for prostate treatment planning.

| Acceptable with minor edits |  |  |  |  |  |  |  |  |  |  |  |  |
| --- | --- | --- | --- | --- | --- | --- | --- | --- | --- | --- | --- | --- |
| Scenario | Prostate | SV | Rectum | Sigmoid | Bladder | FM | PB | LNCTV | BB | Cord | CE | Plan |
| 1 | 100% | 100% | 100% | 93% | 100% | 100% | 100% | 100% | 67% | 100% | 100% | 93% |
| 2 | 100% | 100% | 100% | 100% | 100% | 100% | 100% | 100% | 90% | 100% | 100% | 100% |
| 3 | 100% | 83% | 100% | 100% | 100% | 100% | 100% | 100% | 40% | 100% | 100% | 100% |
| 4 | 100% | 100% | 100% | 100% | 100% | 100% | 100% | 100% | 63% | 100% | 100% | 100% |
| All | **100%** | **98%** | **100%** | **98%** | **100%** | **100%** | **100%** | **100%** | **69%** | **100%** | **100%** | **98%** |
| Use-as-is |  |  |  |  |  |  |  |  |  |  |  |  |
| Scenario | Prostate | SV | Rectum | Sigmoid | Bladder | FM | PB | LNCTV | BB | Cord | CE | Plan |
| 1 | 71% | 86% | 86% | 79% | 93% | 93% | 100% | 100% | 0% | 100% | 100% | 86% |
| 2 | 91% | 91% | 91% | 64% | 100% | 100% | 100% | 100% | 20% | 100% | 100% | 91% |
| 3 | 33% | 0% | 100% | 50% | 100% | 100% | 100% | 100% | 0% | 100% | 100% | 50% |
| 4 | 67% | 67% | 100% | 78% | 78% | 100% | 100% | 100% | 25% | 100% | 100% | 44% |
| All | **70%** | **70%** | **93%** | **70%** | **93%** | **98%** | **100%** | **100%** | **14%** | **100%** | **100%** | **73%** |

Scenario 1,2,3,4 = prostate directed (n = 14), prostate directed + nodes (n = 11), postoperative (n = 6), postoperative + nodes (n = 9), respectively; SV = seminal vesicles; FM = femoral heads; LNCTV = lymph node clinical target volume; BB = bowel bag; CE = cauda equina.

For prostate cancer treatment plans, 23 of 40 test set plans were used for quantitative dose-volume histogram analysis and the percentage of dosimetric criteria (NRG-GU009) passing per patient was calculated across all PTVs and OARs. On average, 77% ± 21% had plans in which all PTV criteria passed per protocol; 91% ±13% of patients had plans in which all PTVs had variation acceptable criteria. For OARs, this was 98% ± 4% and 100% ± 2%, for per protocol and variation acceptable criteria, respectively. The supplementary material contains more information on PTV and OAR dosimetric criteria.

For the end-to-end cervical cancer solution, auto-contours and plans were scored by RTTQA and not edited. Contour quality for cervical cancer targets and OARs was high, with at least 90% of all auto-contours rated to be acceptable without edit (Table 4). All plans and contours were at least acceptable with minor edits. Variation in auto-contouring was mostly seen at cranial and caudal ends of volumes (especially nodal volumes) to a varying degree. However, this appeared less significant when viewed in all 3 planes. In some instances, the nodal CTV and PAN did not completely incorporate the entire vessel circumference on the most superior slices. However, this was often compensated for by adjacent nodal structures where appropriate, and following CTV to PTV expansion, the volumes were felt to be clinically acceptable.

**Table 4.** Physician scores for contour and plan quality for cervical treatment planning.

| Use-as-is (targets and plan) |  |  |  |  |  |  |
| --- | --- | --- | --- | --- | --- | --- |
| Scenario | CTVn | PAN | CTVp | ITV45 | PTV45 | Plan |
| 1 | 100% | N/A | 100% | 100% | 100% | 80% |
| 2 | 100% | 100% | 80% | 100% | 100% | 80% |
| All | 100% | 100% | 90% | 100% | 100% | 80% |

| Use-as-is (organs at risk) |  |  |  |  |  |  |  |  |  |  |
| --- | --- | --- | --- | --- | --- | --- | --- | --- | --- | --- |
| Scenario | Rectum | Bladder | Bowel* | Cord | FM | Kidney | Sacrum | L4 | L5 | Liver |
| 1 | 100% | 100% | 100% | 100% | 100% | 100% | 100% | 80% | 100% | 100% |
| 2 | 100% | 100% | 80% | 100% | 100% | 100% | 100% | 100% | 100% | 100% |
| All | 100% | 100% | 90% | 100% | 100% | 100% | 100% | 90% | 100% | 100% |

**Scenario 1 = PAN excluded (n = 5); Scenario 2 = PAN included (n = 5); Bowel\* = Bowel – CTV; FM= femoral heads.**

All ‘hard dose constraints’ set by EMBRACE II (section 9.9 in EMBRACE II protocol) were met for all targets and OARs. Please see the Supplemental Materia for a table of all criteria. However, for 2/10 patients who had numerous involved nodes close to bowel, bowel dose exceeded D2cc>57.5Gy. For this reason, plan quality score was set at 3 for two patients. One general comment from plan quality review was that the dose to the high dose PTV (treating gross nodal disease) could be tighter but was still acceptable.

## 5. DISCUSSION

The RPA was developed and deployed to improve the availability and quality of radiotherapy in LMICs, consistent with the mission of the NCI Center for Global Health. Through long-term sustained collaborations with cancer experts throughout the world, the RPA was designed to be practical, user-friendly, and cost-free, with a focus on getting treatment to patients that needed them and integrating easily into existing (and varied) clinical workflows. Since FDA clearance in 2023 and first patient treated in 2024, the RPA continues to be used for patient care (with increased uptake throughout 2025). The ongoing use has also highlighted opportunities for improvement, such as the development of full end-to-end radiotherapy contouring and planning in a single step. This work highlights the development and validation of the RPA single step, fully-integrated workflow for prostate and cervical cancer. This single step process operates without intervention (“end-to-end”) by a physician, physicist, or dosimetrist.

The cervical treatment planning solution is compliant with EMBRACE II guidelines, auto-contours 17 structures and produces high-quality treatment plans. Similarly, the prostate treatment planning solution accommodates the prostate directed and prostate bed (with and without nodes) setting, auto-contours 17 structures, and produces high quality-treatment plans which meet NRG-GU009 guidelines.

Our expert evaluation highlights that target auto-contouring required at most minor edits for both the prostate/prostate bed and cervix PTV45 with 70% and 100% of cases requiring no edits for prostate and cervix, respectively. Furthermore, physician reported clinical acceptability of KBP plans indicated that 73% of prostate and 80% of cervical plans did not require edits. In addition, the approach for end-to-end cervical cancer treatment planning is the first which explicitly developed approaches with CT-based EMBRACE II guidelines in mind.

In terms of quantitative accuracy of our approaches for auto-contouring the female and male pelvis compare favorably with the latest approaches in the literature (See Supplementary Material for an in-depth review). The quality of these approaches have been scored by clinical trial experts from the RTTQA group and cancer experts from multiple institutions around the world. They are not focused on suiting the standards of a single institution, but those on an international level, so as to promote a sustainable adoption of AI technology.

One limitation of this work is that CT-only workflows were developed for both cervical and prostate auto-contouring. While MR remains the standard of care for high-resource clinics to contour the targets for the cervix (per EMBRACE II guidelines), CT-based workflows are the most widely accessible [cite]. Furthermore, while the cervix approach in this work fully constructs the PTV from component contours according to EMBRACE II guidelines, it does so with a standard approach, relying on one CT. Thus, our auto-contouring approach is not designed to incorporate or derive margins from multiple CT scans with variable bladder filling and is a topic of future work. This work does not incorporate a means to auto-contour gross nodal disease, and the user must upload these contours upon upload of the CT. Another important point to note, is that while fully-automated treatment planning may have advantages, rigorous review of the auto-contours and plan is essential. As De Kerf et al notes in the work discussing the risks of fully-automated workflows, such workflows introduce new risks and cannot guarantee the best output for every individual [36]. However, this claim may imply that there exists a workflow that can when combined with specialized, automated quality assurance tools. It has been demonstrated, also through failure modes and effects analysis, that there exist opportunities for automated quality assurance to minimize risk as shown by Nealon et al [37]. For the RPA project, this is an ongoing effort for each cancer site which is deployed. It is important to note that these two fully automated workflows were designed for a prospective observational trial (ARCHERY [38]) of AI in which the RPA is used to compare to the manual plan. Valuable insights in safety, efficiency, and workflow will be gained upon its completion and future prospective studies of these prostate and cervix end-to-end solutions are needed.

The RPA was designed to meet a need, namely, staffing and resource shortages. While it is well known that a shortage of approximately 50,000 medical physicists and oncologists exists in radiotherapy [39], the advent of artificial intelligence tools has been hailed as a possible solution to close the resource gap [40]. However, this presents new challenges, as guidance in the commissioning of AI tools is lacking. In addition, there is a lack of studies from LMICs investigating adoption of AI and quantitative accuracy of AI-based tools.

## 6. CONCLUSION

The RPA was created as part of the NCI Center for Global Health ACTs program; version 1 now is being used clinically in South Africa to treat patients with cancers of the cervix, post-mastectomy breast, and head/neck. The opportunity to further refine the quality and efficiency of this process is based on ongoing close collaborations with cancer experts in LMICs, and the potential to create validated end-to-end processes for VMAT treatment planning for prostate and cervical cancer patients was a clear next step. In this work, AI-powered auto-contouring produced clinically acceptable results for both cancer sites, and knowledge-based planning was leveraged to produce treatment plans that require no edits for 73-80% of cases in the retrospective setting. Our global collaborations are crucial to address the “tidal wave” of cancers throughout the world, and the RPA will continue to integrate improvements in quality and efficiency to address this need.

## Data Availability

patient data are not available at this time

## Acknowledgements

This work was supported in part through the National Institute of Health, National Cancer Institute (Award No. UH2CA202665). This work was supported in part from the National Science Foundation Graduate Research Fellowship Program under Grant No. 2043424. This work was in part supported through the Rising Tide Foundation (Award No. CCR-21-300, ARCHERY study). This work was funded in part by Varian Medical Systems. We would like to acknowledge the support of Tumor Measurement Initiative (TMI) through the University of Texas MD Anderson STrategic Initiative DEvelopment Program (STRIDE). We thank those from the Radiotherapy Trials Quality Assurance (RTTQA) group for their time and expertise. Author TJ Netherton would like to acknowledge the support of the Loan Repayment Award from the National Institute of Health, National Cancer Institute. The National Radiotherapy Trials Quality Assurance (RTTQA) Group is funded by the National Institute for Health and Care Research (NIHR).

## Notes

### Competing Interest Statement

The authors have declared no competing interest.

### Author Declarations

This study was conducted under a protocol which was approved by the University of Texas MD Anderson Cancer Center Institutional Review Board

