## supplemental document for "AI-Powered Radiotherapy for Resource-Limited Settings: Advancing Cervical and Prostate Cancer Treatment Planning with the Radiation Planning Assistant (RPA)"

**Supplemental Material**

**Dataset and study design diagrams**

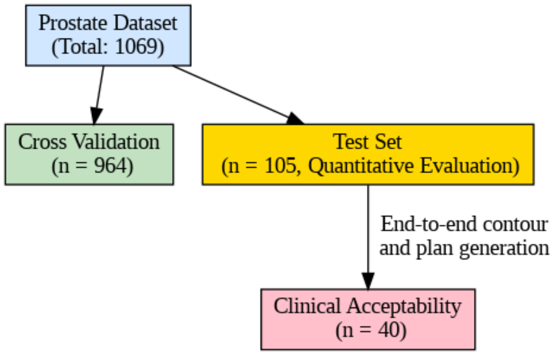

Figure 1. Prostate dataset diagram

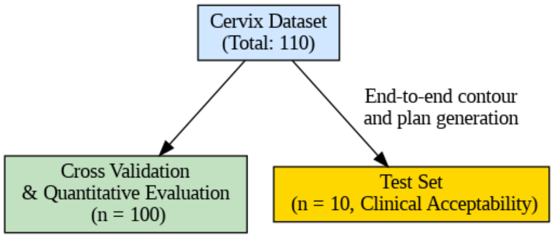

Figure 2. Cervical dataset diagram

**Curation of ground truth contours for prostate**

The MDA cohort was evaluated to ensure that CTs, manual contours and VMAT treatment plans were clinically approved and used in treatment. Each contour selected for training was individually evaluated for observance to guidelines for model training with supervision from a radiation oncologist (see Table 1). Selected clinical contours include Prostate, Prostate Bed, Seminal Vesicles (SV), SV-Fossa, Rectum, Sigmoid, Bladder, Femoral Heads, spaceoar Hydrogel and Rectal Balloon. Not all patients had each of the 15 contours present in the MDA autocontouring cohort. To create a full contour set per patient, a nnU-Net model [1] was trained for each of the contours. Missing contours on each patient from the MDA cohort were then filled in from these model predictions.

| **Organization** | **Structures** | **Reference** |
|---|---|---|
| ESTRO | Intact Prostate | *ESTRO ACROP consensus guideline on CT- and MRI-based target volume delineation for primary radiation therapy of localized prostate cancer* [2] |

| | | |
|---|---|---|
| RTOG | Post-operative Prostate, Normal Pelvic Tissues | • *Development of RTOG consensus guidelines for the definition of the clinical target volume for postoperative conformal radiation therapy for prostate cancer* [3]<br>• *Pelvic Normal Tissue Contouring Guidelines for Radiation Therapy: A Radiation Therapy Oncology Group Consensus Panel Atlas* [4] |
| NRG | Pelvic Lymph Nodes | *NRG Oncology Updated International Consensus Atlas on Pelvic Lymph Node Volumes for Intact and Postoperative Prostate Cancer* [5] |

*Table 1 Summary of contouring guidelines observed for curation of ground truth data for deep learning model development.*

**Post processing for prostate auto-contours**

To combine all the predicted structures from the 5 models, additional contour corrections were necessary. Overlap between predicted structures needed to be corrected between targets and OARs and physician input was crucial to this process. The priority of overlap was set to override as follows: prostate, seminal vesicles, rectum, sigmoid, bladder, lymph node CTV, femoral heads, penile bulb, bowel bag, cauda equina, spinal cord and kidneys. For continuous structures such as Prostate Bed/SV-fossa, Rectum/Sigmoid, and Cauda Equina/Spinal Cord), the boundaries required adjustment to avoid multiple predictions on same slice axially (see Figure ). Additionally for prostate, a 1.5mm and 1mm anterior and lateral reduction was necessary to avoid contouring into the neurovascular bundle surrounding the prostate. For prostate bed, a similar anterior and superior 1mm and 2mm reduction was required to avoid excessive contouring into the bladder. For each structure pair, the middle slice in which there was overlap was chosen as the boundary between ROIs. As total seminal vesicle volume was contoured, for intact treatment cases, the seminal vesicles are truncated based on desired treatment extent. For example, if the physician indicates that they want to treat the proximal 1.5cm, then the seminal vesicles are truncated at the 1.5cm point.

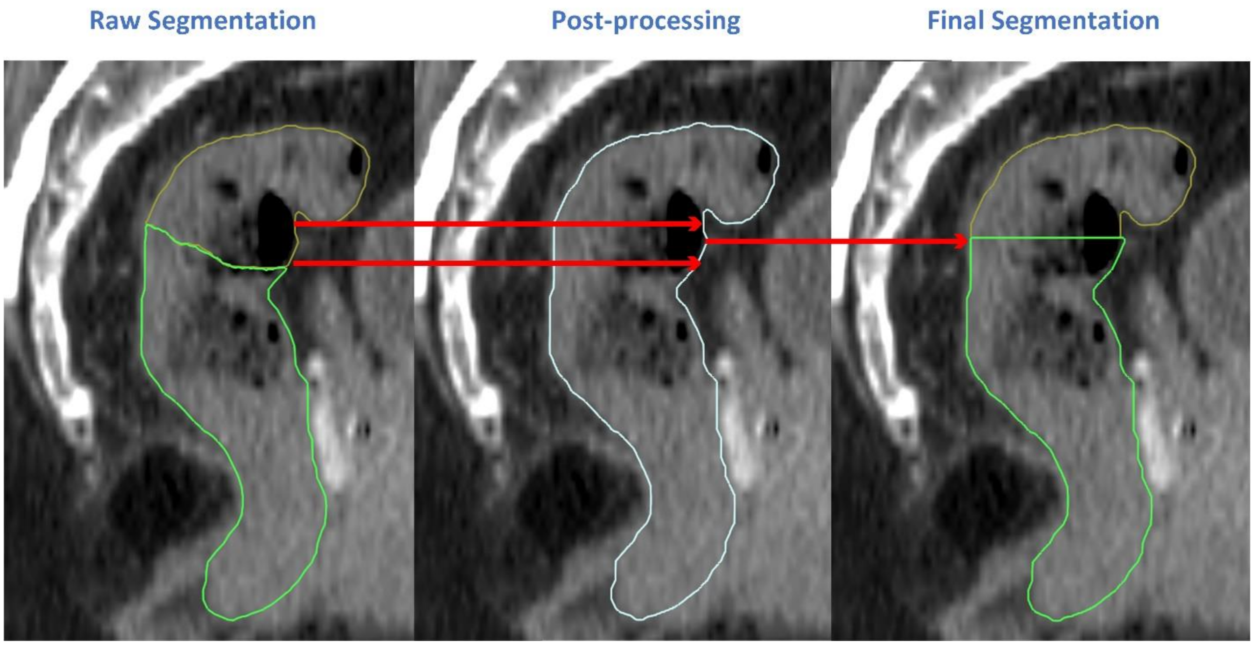

*Figure 3 Boundary correction performed on generated segmentations of model predictions. An example of an auto-contoured rectum and sigmoid prediction being reset to the median overlapping slice from the most inferior and superior borders of the contour boundaries.*

**Scoring Rubric for physician review**

| **Score** | **Acceptability** | **Description** |
|---|---|---|
| 5 | Acceptable, use as is | Clinically acceptable, could be used for treatment without change |
| 4 | Acceptable, minor edits that are not necessary | Stylistic differences, but not clinically important; the current contours/plans are acceptable |
| 3 | Acceptable after redress, minor edits that are necessary | Edits that are clinically important, but it is more efficient to edit the automatically generated contours/plans than to start from scratch |
| 2 | Unacceptable, major edits | Edits that are required to ensure appropriate treatment and sufficiently significant that the user would prefer to start from scratch |
| 1 | Unacceptable, unusable | Automatically generated contours/plans are so bad that they are unusable (i.e., wrong body area, outside confines of body, etc.) |

*Table 1 The table details the 5-point scale for evaluating the quality of generated auto-contours and auto-plans.*

**Prostate Physician Review**

| ID | Prostate | Seminal Vesicle | Rectum | Sigmoid | Bladder | Femoral Heads | Penile Bulb | Lymph Node CTV | Bowel Bag | SpinalCord | Cauda-Equina | spaceoar | Plan | Case |
|---|---|---|---|---|---|---|---|---|---|---|---|---|---|---|
| pros1249 | 5 | 5 | 4 | 4 | 4 | 3 | 5 | 5 | 3 | na | na | na | 5 | intact |
| pros1266 | 3 | 3 | 4 | 1 | 5 | 5 | 5 | 5 | 2 | na | na | na | 3 | intact |
| pros1270 | 3 | 3 | 4 | 5 | 5 | 5 | 5 | 5 | 3 | na | na | na | 4 | intact |
| pros2199 | 5 | 5 | 3 | 3 | 3 | 5 | 5 | na | na | na | na | na | 5 | intact |
| pros2200 | 4 | 4 | 3 | 3 | 5 | 4 | 5 | na | na | na | na | na | 4 | intact |
| pros2135 | 4 | 5 | 5 | 3 | 5 | 5 | 5 | 5 | 3 | na | na | na | 5 | intact+node |
| pros2185 | 5 | 5 | 5 | 3 | 5 | 4 | 5 | 5 | 3 | na | na | na | 5 | intact+node |
| pros2186 | 5 | 5 | 4 | 5 | 5 | 4 | 5 | 5 | 3 | na | na | na | 3 | intact+node |
| pros2104 | 3 | 3 | 5 | 4 | 5 | 5 | 5 | na | na | na | na | na | 3 | postop |
| pros1190 | 3 | 3 | 4 | 5 | 3 | 5 | 5 | 4 | 3 | na | na | na | 3 | postop+node |
| pros2157 | 3 | 3 | 4 | 3 | 3 | 5 | 5 | 5 | na | na | na | na | 3 | postop+node |
| pros1181 | 4 | 4 | 5 | 3 | 5 | 5 | 5 | 4 | 2 | 5 | 5 | 5 | 5 | intact+node |
| pros1240 | 3 | 3 | 5 | 3 | 5 | 5 | 4 | na | 2 | na | na | na | 3 | postop |
| pros1245 | 3 | 3 | 5 | 3 | 5 | 5 | 5 | 4 | 2 | na | na | na | 4 | postop |
| pros1274 | 3 | 2 | 5 | 3 | 5 | 5 | 5 | 4 | 2 | 5 | 5 | na | 3 | postop |
| pros1199 | 4 | 4 | 5 | 3 | 5 | 5 | 5 | 5 | 2 | 5 | 5 | na | 4 | postop+node |
| pros1206 | 4 | 4 | 5 | 5 | 4 | 5 | 4 | 4 | 2 | 5 | 5 | na | 5 | intact |
| pros1220 | 3 | 5 | 5 | 5 | 5 | 5 | 5 | 4 | 3 | 5 | 5 | na | 5 | intact |
| pros2101 | 4 | 4 | 4 | 4 | 5 | 5 | 4 | na | na | na | na | na | 5 | intact |
| pros2105 | 4 | 5 | 5 | 4 | 5 | 5 | 4 | na | 3 | na | na | na | 2 | intact |
| pros2148 | 4 | 5 | 5 | 5 | 4 | 4 | 4 | na | na | na | na | na | 4 | intact |
| pros2150 | 4 | 5 | 5 | 4 | 5 | 5 | 4 | na | na | na | na | na | 5 | intact |
| pros2152 | 3 | 4 | 4 | 5 | 5 | 5 | 4 | na | na | na | na | na | 5 | intact |
| pros2198 | 4 | 4 | 4 | 4 | 4 | 5 | 4 | na | na | na | na | na | 5 | intact |

| | | | | | | | | | | | | | |
|---|---|---|---|---|---|---|---|---|---|---|---|---|---|
| pros2210 | 4 | 4 | 5 | 5 | 5 | 5 | 4 | na | na | na | na | na | 5 | intact |
| pros1186 | 4 | 5 | 5 | 5 | 5 | 5 | 4 | 5 | 3 | 5 | 5 | na | 4 | intact+node |
| pros2103 | 4 | 5 | 5 | 4 | 5 | 5 | 4 | 5 | 3 | 5 | na | na | 4 | intact+node |
| pros2143 | 3 | 3 | 3 | 4 | 4 | 5 | 4 | 4 | 3 | 5 | na | na | 4 | intact+node |
| pros2162 | 4 | 4 | 5 | 5 | 5 | 4 | 4 | 4 | 4 | na | na | na | 4 | intact+node |
| pros2163 | 5 | 5 | 4 | 3 | 5 | 5 | 4 | 4 | 3 | na | na | na | 4 | intact+node |
| pros2183 | 4 | 5 | 5 | 5 | 5 | 5 | 4 | 5 | 4 | na | na | na | 4 | intact+node |
| pros2197 | 4 | 4 | 5 | 5 | 5 | 5 | 4 | 4 | na | na | na | na | 4 | intact+node |
| pros1207 | 4 | 3 | 5 | 5 | 4 | 5 | 4 | 4 | 3 | 5 | 5 | na | 5 | postop |
| pros1226 | 5 | 3 | 5 | 5 | 5 | 5 | 5 | 5 | 3 | 5 | 5 | na | 5 | postop |
| pros1190 | 5 | 4 | 5 | 5 | 4 | 5 | 4 | 4 | 2 | na | na | na | 4 | postop+node |
| pros1198 | 4 | 4 | 5 | 5 | 5 | 5 | 4 | 4 | 3 | na | na | na | 3 | postop+node |
| pros1201 | 4 | 4 | 4 | 4 | 4 | 5 | 5 | 5 | 2 | 5 | 5 | na | 4 | postop+node |
| pros2102 | 3 | 3 | 4 | 4 | 5 | 4 | 5 | 4 | 3 | 5 | na | na | 3 | postop+node |
| pros2108 | 4 | 4 | 5 | 5 | 5 | 5 | 4 | 5 | 4 | na | na | na | 5 | postop+node |
| pros2141 | 4 | 4 | 4 | 4 | 5 | 5 | 5 | 4 | 4 | na | na | na | 3 | postop+node |

**Prostate**

| ID | PTV PP | PTV VA | OAR PP | OAR VA | Plan Quality | Case |
|---|---|---|---|---|---|---|
| pros2101 | 67% | 100% | 100% | 100% | 5 | intact |
| pros2105 | 63% | 88% | 100% | 100% | 2 | intact |
| pros2148 | 100% | 100% | 100% | 100% | 4 | intact |
| pros2150 | 100% | 100% | 100% | 100% | 5 | intact |
| pros2152 | 100% | 100% | 100% | 100% | 5 | intact |
| pros2198 | 100% | 100% | 100% | 100% | 5 | intact |

| | | | | | | |
|---|---|---|---|---|---|---|
| pros2199 | 100% | 100% | 100% | 100% | 5 | intact |
| pros2200 | 100% | 100% | 91% | 100% | 4 | intact |
| pros2210 | 100% | 100% | 100% | 100% | 5 | intact |
| pros2103 | 60% | 80% | 100% | 100% | 4 | intact+node |
| pros2135 | 40% | 80% | 100% | 100% | 5 | intact+node |
| pros2143 | 80% | 100% | 100% | 100% | 4 | intact+node |
| pros2162 | 80% | 100% | 100% | 100% | 4 | intact+node |
| pros2163 | 60% | 100% | 100% | 100% | 4 | intact+node |
| pros2183 | 80% | 100% | 100% | 100% | 4 | intact+node |
| pros2185 | 50% | 75% | 100% | 100% | 5 | intact+node |
| pros2186 | 80% | 80% | 100% | 100% | 3 | intact+node |
| pros2197 | 100% | 100% | 100% | 100% | 4 | intact+node |
| pros2104 | 67% | 100% | 91% | 100% | 3 | postop |
| pros2102 | 80% | 80% | 100% | 100% | 3 | postop+node |
| pros2108 | 80% | 80% | 83% | 91% | 5 | postop+node |
| pros2141 | 50% | 75% | 100% | 100% | 3 | postop+node |
| pros2157 | 38% | 50% | 100% | 100% | 3 | postop+node |

**PP = per protocol, VA = variation acceptable**

**Cervix Physician Review**

| De-identified ID | CTVn | PAN | CTVp | ITV45 | PTV45 | Rectum | Bladder | Bowel - CTV | Spinal Cord | Femur_L | Femur_R | Kidney_L | Kidney_R | Sacrum | L4 | L5 | Liver | Body | Plan Quality |
|---|---|---|---|---|---|---|---|---|---|---|---|---|---|---|---|---|---|---|---|
| RxP_GYN002 | 4 | NA | 4 | 5 | 5 | 5 | 5 | 5 | 5 | 5 | 5 | 5 | 5 | 4 | 5 | 5 | 5 | 5 | 4 |
| RxP_GYN017 | 5 | NA | 5 | 5 | 5 | 5 | 5 | 4 | 5 | 5 | 5 | 5 | 5 | 4 | 5 | 5 | 4 | 5 | 4 |

| | | | | | | | | | | | | | | | | | | | |
|---|---|---|---|---|---|---|---|---|---|---|---|---|---|---|---|---|---|---|---|
| RxP_GYN021 | 4 | NA | 4 | 5 | 5 | 5 | 5 | 4 | 5 | 5 | 5 | 5 | 5 | 4 | 5 | 5 | 5 | 5 | 3 |
| RxP_GYN023 | 4 | NA | 5 | 4 | 5 | 5 | 5 | 4 | 5 | 4 | 4 | 5 | 5 | 5 | 3 | 5 | 5 | 5 | 4 |
| RxP_GYN029 | 4 | NA | 4 | 5 | 5 | 4 | 5 | 4 | 5 | 4 | 4 | 5 | 5 | 4 | 5 | 5 | 5 | 5 | 4 |
| RxP_GYN005 | 5 | 4 | 4 | 5 | 5 | 4 | 4 | 4 | 5 | 5 | 5 | 5 | 4 | 5 | 5 | 5 | 4 | 5 | 3 |
| RxP_GYN015 | 5 | 4 | 3 | 4 | 5 | 4 | 4 | 4 | 5 | 4 | 4 | 5 | 5 | 4 | 5 | 5 | 4 | 5 | 4 |
| RxP_GYN018 | 4 | 4 | 4 | 5 | 4 | 4 | 5 | 3 | 5 | 5 | 5 | 5 | 5 | 4 | 5 | 5 | 4 | 5 | 4 |
| RxP_GYN022 | 5 | 4 | 5 | 4 | 5 | 4 | 5 | 5 | 5 | 5 | 4 | 5 | 5 | 4 | 5 | 5 | 5 | 5 | 5 |
| RxP_GYN027 | 4 | 4 | 4 | 4 | 5 | 4 | 5 | 4 | 5 | 4 | 4 | 5 | 5 | 4 | 5 | 5 | 5 | 5 | 4 |

**Cervix Dosimetric Scoring**

| | | D98%>51.75<br>D2cc < 61.53 | D98%>57.5 | Dmin<br>>42.75 | V42.75<br>> 95%<br>D2cc<br><48.15*<br>*not boost | Dmax<47.3*<br><br>Dmax<60.4<br>*not boost | Dmax<47.3*<br><br>Dmax<60.4<br>*not boost | Dmax<47.3*<br><br>Dmax<60.4<br>*not boost | Dmax < 48 | Dmax < 50 | Dmean < 15 | Overall Plan qualiy Score 1-5 | |
|---|---|---|---|---|---|---|---|---|---|---|---|---|---|
| **De-identified ID** | **PAN treated** | **PTV-N** | **GTV-N** | **ITV45** | **PTV45** | **Rectum** | **Bladder** | **Bowel - CTV** | **Spinal Cord** | **Femurs** | **Kidney** | **Plan** | **Comment** |

| | | | | | | | | | | | | | |
|---|---|---|---|---|---|---|---|---|---|---|---|---|---|
| **RxP_GYN0 02** | No | 56.72;59.24 | not evaluated | 42.93 | 99.9 | 46.5 | 46.8 | 54.1 | 25.12 | 32.8 | N/A | 4 | PTV5750 might be tighter |
| **RxP_GYN0 17** | No | 57.35;59.03 | not evaluated | 44.235 | 99.9 | 46.28 | 46.55 | 54.15 | 16.53 | 31.37 | N/A | 4 | PTV5750 might be tighter |
| **RxP_GYN0 21** | No | 56.56;59.18 | not evaluated | 42.975 | 99.8 | 46.59 | 46.94 | 58.01 | 18.23 | 27.88 | 0.44 | 3 | bowel out of tolerance, |
| **RxP_GYN0 23** | No | 56.87;59 | not evaluated | 43.38 | 99.9 | 46.29 | 46.69 | 47.22 | 23.94 | 30.42 | N/A | 4 | PTV5750 might be tighter |
| **RxP_GYN0 29** | No | 56.7;59.1 | not evaluated | 44.68 | 99.9 | 46.47 | 46.7 | 46.9 | 27.4 | 31.4 | N/A | 4 | PTV5750 might be tighter |
| **RxP_GYN0 05** | Yes | 55.65;59.62 | not evaluated | 42.525 | 99 | 46.79 | 47.87 | 58.33 | 33.98 | 28.56 | 11.42 | 3 | bowel out of tolerance, |
| **RxP_GYN0 15** | Yes | 55.94;59.33 | not evaluated | 43.11 | 99.1 | 46.62 | 47.07 | 52.39 | 35.31 | 31.66 | 12.27 | 4 | PTV5750 might be tighter |
| **RxP_GYN0 18** | Yes | 56.37;59.26 | not evaluated | 43.47 | 99.7 | 46.59 | 46.92 | 53.88 | 32.47 | 28.91 | 11.25 | 4 | PTV5750 might be tighter |
| **RxP_GYN0 22** | Yes | N/A | not evaluated | 44.01 | 99.8 | 46.34 | 46.44 | 46.6 | 35.12 | 28.7 | 13.73 | 5 | |
| **RxP_GYN0 27** | Yes | 99.3;59.6 | not evaluated | 44.6 | 99.3 | 46.7 | 54.4 | 57.1 | 32.9 | 27.7 | 11.9 | 4 | PTV5750 might be tighter |

**Prostate Auto-contouring Literature Review Table**

In regard to current advances in prostate auto-contouring, typical training datasets size can range between 47 (Kawula et al., 2022) and 1530 patients (Polymeri et al., 2024). Dice scores can vary widely, but approach 0.92 in advanced models like Arjmandi et al. (2024). With deep learning, more consistent and higher accuracy was achieved—Wen et al. (2024) reported CTV Dice scores up to 0.86, while Balagopal et al. (2021) showed personalized, style-aware improvements using PSA-Net. Clinical acceptability for CTVs, where available, ranged from 60.9% to 87%, reflecting both methodological advances and continued challenges in complex anatomical regions. To our knowledge, our work is the first which can auto-contour and auto-plan end-to-end in both the prostate directed and prostate bed setting for prostate carcinoma.

For cervical cancer CTVs specifically, studies demonstrated a wide range of training dataset sizes (64–272 patients), Dice scores (0.70–0.89), and clinical acceptability (63.5–98%). Early CTV segmentation efforts, such as Rhee DJ et al. (2020), achieved moderate Dice scores (0.76–0.86) but highlighted challenges in nodal CTV delineation. Later advancements, like the TCAS network (Ma C ying et al., 2022), improved CTV Dice scores to 0.89 through multi-channel architectures. However, lower scores persisted for parametrial areas (e.g., dCTV2: 0.70) due to anatomical variability. Clinical acceptability for CTVs remain variable, with studies like Zhang C et al. (2022) reporting 91.5% acceptance, while others emphasized the need for manual edits. The approach for end-to-end cervical cancer treatment planning developed in this work achieves excellent auto-contouring and autoplanning performance. To our knowledge, it is the first which explicitly developed approaches with CT-based EMBRACE II guidelines in mind.

| Title | Auto-contouring Method | # Structures | Target & OAR | Patients (Training/Testing) | Dice Scor | Physician Clinical Acceptability |
|---|---|---|---|---|---|---|
| Evaluation of the accuracy of automated segmentation based on deep learning for prostate cancer patients. [6]<br><br>Miura et al. (2025) | MVision AI Contour | 7 | Targets: Prostate, Seminal Vesicles (SV)<br><br>OAR: Bladder, Rectum, Left Femoral Head, Right Femoral Head, Penile Bulb | Test: 10 patients<br><br>Train: N/A | Prostate = 0.86, Seminal Vesicles (SV) = 0.80, Bladder = 0.96, Rectum = 0.92, Left Femoral Head = 0.97, Right Femoral Head = 0.97, Penile Bulb = 0.64 | N/A |
| Deep learning based clinical target volumes contouring for prostate cancer: Easy and efficient application.[7] Wen et al. (2024) | DeepLabV3+, UNet++, and 3D U-Net | 2 | Targets:<br><br>1-CTVn (pelvic lymph nodes)<br><br>2-CTVp (prostate tumors or prostate tumor beds) | Train: 167 patients (135 for training, 32 for validation) | Radical Radiotherapy Model:<br><br>CTVn = 0.85<br><br>CTVp = 0.84<br><br>Postoperative Radiotherapy Model: | 82% |

| | | | | | | |
|---|---|---|---|---|---|---|
| | | | OARs: N/A | Test: 30 patients | CTVn = 0.86<br>CTVp = 0.79 | |
| Artificial Intelligence-Based Organ Delineation for Radiation Treatment Planning of Prostate Cancer on Computed Tomography.[8] Polymeri et al. (2024) | 3D U-Net fully convolutional neural network | 3 | Target: Prostate (with and without seminal vesicles)<br><br>OARs: Urinary Bladder, Rectum | Train & Validation: ~75% of 1530 patients (approx. 1147)<br><br>Test: ~25% of 1530 patients (approx. 383 patients) | Prostate only = 0.82<br>Prostate + seminal vesicles = 0.83<br>Urinary Bladder = 0.95<br>Rectum = 0.88 | N/A |
| Automated contouring of CTV and OARs in planning CT scans using novel hybrid convolution-transformer networks for prostate cancer radiotherapy. [9]Arjmandi et al. (2024) | Vision Transformer (CNN-ViT), specifically VGG16-UNet-ViT and ResNet50-UNet-ViT architectures. | 5 | Target: Prostate (CTV)<br><br>OARs: Bladder, Rectum, Right Femoral Head (RFH), Left Femoral Head (LFH) | 104 patients (70% train, 10% validation, 20% test). Approximately 73 training: 10 validation: 21 testing cases. | Prostate = 91.75<br>Bladder = 95.32<br>Rectum = 87.00<br>RFH = 96.30<br>LFH = 96.34 | N/A |
| Real-world validation of Artificial Intelligence-based Computed Tomography auto-contouring for prostate cancer radiotherapy planning.[10] | U-Net | 5 | Targets (CTV): Prostate, Seminal Vesicles | 20 patients (all used for validation) | Prostate = 0.86<br>Seminal Vesicles = 0.76 | Median clinical score of 4/5 (little editing needed |

| | | | | | | |
|---|---|---|---|---|---|---|
| Palazzo et al., (2023) | | | OARs: Bladder, Rectum, Femoral Heads + Femurs | | Bladder = 0.95<br>Rectum = 0.84<br>Femoral Heads + Femurs = 0.83 | |
| Dosimetric impact of deep learning-based CT auto-segmentation on radiation therapy treatment planning for prostate cancer.<br>Kawula et al.[11] (2022) | 3D U-Net based on V-Net architecture | 3 | Target: Prostate<br>OARs: Bladder, Rectum | Training: 47<br>Validation: 11<br>Test: 11 | Prostate: 0.87 ± 0.03<br>Bladder: 0.97 ± 0.01<br>Rectum: 0.89 ± 0.04<br>Total = 0.91 | 91% |
| Evaluating the clinical acceptability of deep learning contours of prostate and organs-at-risk in an automated prostate treatment planning process.[12]<br>Duan et al. (2022) | 3D U-Net | 7 | Target: Prostate<br>OARs: Bladder, Rectum, Left Femoral Head, Right Femoral Head, Penile Bulb, Seminal Vesicles (SVs) | Training/Validation: 84<br>Testing: 23 | Prostate: 0.83 ± 0.05<br>Bladder: 0.93 ± 0.04<br>Rectum: 0.85 ± 0.05<br>Seminal Vesicles: 0.72 ± 0.1<br>Femoral Heads: 0.96 ± 0.01 (left), 0.97 ± 0.01 (right)<br>Penile Bulb: 0.53 ± 0.17 | (60.9%) |
| Automated contour propagation of the prostate from pCT to CBCT images via deep unsupervised learning.[13] Liang et al. (2021) | 3D Attention U-Net | 1 | Target: Prostate<br>OARs: N/A | Training: 180<br>Validation: 12<br>Testing: 59 | Prostate:<br>Group 1: 0.83 ± 0.04<br>Group 2 (Consensus): 0.85 ± 0.04 | N/A |
| PSA-Net: Deep learning–based physician style–aware segmentation network for | PSA-Net | 3 | Target: Clinical Target Volume (CTV) for | UT Southwestern Dataset: | CTV Dice Scores (UTSW):<br>Mixed-Style Model: 87.8%, 82.8%, | 87% |

| | | | | | | |
|---|---|---|---|---|---|---|
| postoperative prostate cancer clinical target volumes.[14]<br>Balagopal et al. 2021 | | | Postoperative Prostate Cancer<br>OARs (Supporting Structures): Bladder, Rectum | 373 patients total<br>60 patients for testing (15 per physician)<br>Mayo Clinic Dataset (External Validation): 83 patients<br>53 for training, 30 for testing (for institutional style adaptation) | 83.3%, 80.0% for Physicians 1–4 respectively<br>PSA-Net (Style-Aware): +3.4% average improvement over mixed-style<br>CTV Dice Scores (Mayo):<br>General UTSW Model on Mayo Data: 70.5% (±6.4)<br>Style-Adapted Model (Mayo): +5% improvement over general UTSW model<br>Bladder (Mayo): 94.4% (±3.5)<br>Rectum (Mayo): 87.5% (±6.2) | |
| Comparison of atlas-based auto-segmentation accuracy for radiotherapy in prostate cancer. Aoyama et al. (2021)[15] | Atlas-Based Auto-Segmentation (ABS) | 7 | Targets: Prostate, Seminal Vesicles<br>OARs: Rectum, Bladder, Pubis, Ischium, Femoral Head | Total Patients: 30<br>Training (Atlas Creation): 20 patients<br>Testing: 10 patients | Prostate:<br>sSM: 0.64 (0.27–0.71)<br>sMM: 0.81 (0.66–0.91), $p < 0.01$<br>Seminal Vesicles:<br>sSM: 0.18 (0.01–0.60) | N/A |

| | | | | | | |
|---|---|---|---|---|---|---|
| | | | | | sMM: 0.49 (0.31–0.80), $p < 0.05$<br>Rectum:<br>sSM: 0.57 (0.31–0.77)<br>sMM: 0.81 (0.37–0.91), $p < 0.01$ | |
| A Deep Learning-Based Automated CT Segmentation of Prostate Cancer Anatomy for Radiation Therapy Planning-A Retrospective Multicenter Study.[16] Kiljunen et al. (2020) | Commercial DL-based AST (Automated Segmentation Tool) using a 3D U-Net | 7 | Targets: Prostate, Seminal Vesicles, Lymph Nodes<br>OARs: Bladder, Rectum, Femoral Heads, Penile Bulb | Training :900 patients<br>Evaluation: 30 patients | Prostate: 0.82<br>Seminal Vesicles: 0.72<br>Bladder: 0.93<br>Rectum: 0.84<br>Femoral Heads: 0.68–0.69<br>Penile Bulb: 0.51<br>Lymph Nodes: 0.80 (in one clinic | N/A |
| Automatic Segmentation of the Prostate on CT Images Using Deep Neural Networks(DNN).[17] Liu et al. (2019) | Deep Neural Network (DNN) | 1 | Target: Prostate<br>OARs:N/A | Group A:<br>Training: 771 patients<br>Validation: 193 patients<br>Testing: 140 patients<br>Group B:<br>10 patients | Group A (Test): 0.85 ± 0.06<br>Group B (Average of 5 Observers): 0.85 ± 0.04<br>Group B (Consensus): 0.88 ± 0.03 | N/A |

| | | | | | | |
|---|---|---|---|---|---|---|
| Comparison of Automated Atlas-Based Segmentation Software for Postoperative Prostate Cancer Radiotherapy. [18]Delpon et al. (2016) | Atlas-Based Segmentation (ABS), | 4 | Target: Prostate Bed (CTV)<br><br>OARs: Bladder, Rectum, Femoral Heads | Training (Atlas Creation): 10 patients<br><br>Testing (Evaluation): 10 patients | emoral Heads (Left): 0.89 – 0.91 (SPICE: 0.70)<br>Femoral Heads (Right): 0.91 – 0.92 (SPICE: 0.72)<br>Bladder: 0.76 – 0.81 (RS: 0.59)<br>Rectum: 0.68 – 0.75 (RS: 0.49)<br>Prostate Bed CTV: 0.37 – 0.67 | N/A |
| Evaluation of atlas-based auto-segmentation software in prostate cancer patients. [19]Greenham et al. (2014) | Atlas-Based Auto-Segmentation (ABAS) | 6 | Target: Prostate<br><br>OARs: Bladder, Rectum, Left Femoral Head, Right Femoral Head, External Patient Contour | Training (Atlas Creation): 10 patients<br><br>Testing (Evaluation Dataset): 24 patients | Bladder DSC (r): 0.95 – 0.99<br>Rectum DSC (r): 0.68 – 0.74<br>Prostate DSC (r): 0.40<br>Right Femoral Head DSC (r): 0.28 – 0.83<br>Left Femoral Head DSC (r): 0.56 – 0.90<br>External Patient Contour (r): 0.61 – 1.00 | Bladder: 77.52% acceptable or minor edits required<br>Rectum: 48.76% acceptable or minor edits required<br><br>Prostate: 21.01% acceptable or minor edits required<br>Right Femoral Head: 94.03% acceptable or |

| | | | | | | |
|---|---|---|---|---|---|---|
| | | | | | | minor edits required<br>Left Femoral Head: 93.98% acceptable or minor edits required<br>External Patient Contour: 76.74% acceptable or minor edits required |
| Segmenting CT Prostate Images Using Population and Patient-Specific Statistics for Radiotherapy[20]. Feng et al. (2009) | Deformable Model using SIFT features | 1 | Target: Prostate<br><br>OAR:N/A | Test: 264<br>Train: 24 | Average DSC: 90.5% ± 4.0<br>Min DSC: 55.1%<br>Median DSC: 91.2%<br>Max DSC: 96.9% | N/A |
| Assessment of Accuracy and Efficiency of Atlas-Based Autosegmentation for Prostate Radiotherapy in a Variety of Clinical Conditions. Simmat et al. [21](2012) | Atlas-Based Autosegmentation (ABAS) | 3 | Target: Prostate<br><br>OAR: Rectum, Bladde | Planning CT: 20 patients<br><br>CBCT (Intra-patient Atlas): 10 patients | Prostate:<br>ABAS: 0.71 ± 0.14<br>iPlan: 0.57 ± 0.19<br>Rectum:<br>ABAS: 0.78 ± 0.11<br>iPlan: 0.84 ± 0.08<br>Bladder:<br>ABAS: 0.86 ± 0.17<br>iPlan: 0.51 ± 0.30 | 90% |

| | | | | | |
|---|---|---|---|---|---|
| | | | | | CBCT Prostate DSC: 0.34 |

**Cervix Auto-contouring literature review Table**

| Title | Ground truth follows EMBRACE II? | Deep Learning Method | Number of Structures | Target & OAR | Patients (Training/Testing) | Dice Score | Physician Clinical Acceptability |
|---|---|---|---|---|---|---|---|
| Development and validation of a deep reinforcement learning algorithm for auto-delineation of organs at risk in cervical cancer radiotherapy [1] Yucheng et al 2025. | Clinical experience | DRL + SAM | 10 | Target: Cervical cancer OARs: Left kidney, Right kidney, Liver, Bladder, Spleen, Duodenum, Bone marrow, Pancreas, Stomach, Rectum | Train: 122; Test: 28 | - Left Kidney: 0.97<br>- Right Kidney: 0.97<br>- Liver: 0.96<br>- Bladder: 0.92<br>- Spleen: 0.94<br>- Duodenum: 0.77<br>- Bone Marrow: 0.84<br>- Pancreas: 0.83<br>- Stomach: 0.92<br>- Rectum: 0.82 | N/A |
| Clinical target volume (CTV) automatic delineation using deep learning network for cervical cancer radiotherapy: | Target based on RTOG | ResCANet | 1 | Target: CTV | 189 training / 47 internal validations + 54 external cervical cancer + 42 endometrial cancer | Internal: 0.748<br>Cervical: 0.734<br>Endometrial: 0.771 | 85% |

| | | | | | | | |
|---|---|---|---|---|---|---|---|
| A study with external validation [2] Wu et al 2025 | | | | | | | |
| Automatic segmentation of high-risk clinical target volume and organs at risk in brachytherapy of cervical cancer with a convolutional neural network [3]<br>Zhu J et al 2024 | Consensus guidelines IBS-GEC_ESTRO-ABS | SERes-U-Net (vs. Res-U-Net and U-Net) | 5 | Target: HR-CTV<br>OARs: Bladder, Rectum, Sigmoid, Bowel Loops | 68 (Training)<br>15 (Testing) | HR-CTV: 0.808<br>Bladder: 0.919<br>Rectum: 0.852<br>Sigmoid: 0.604<br>Bowel: 0.828 | HR-CTV:<br>- 99.3% accepted by Oncologist A<br>- 100% accepted by Oncologist B<br>OARs: Most clinically acceptable (except 25% sigmoid needing revision per Oncologist A) |
| Segmentation of Clinical Target Volume From CT Images for Cervical Cancer Using Deep Learning Images[4]<br>Huang M et al 2023 | N/A | Mnet_IM | 1 | Target: CTV | Train: 164; Val: 35; Test: 36 | VD: 0.8828 | N/A |
| Self-configuring nnU-Net for automatic delineation of the organs at risk and target in high-dose rate cervical brachytherapy, | Consensus guidelines IBS-GEC_ESTRO-ABS | nnU-Net (3DFR) | 3 | Target: HR CTV<br>OARs: Bladder, Rectum | Train: 80; Test: 20 | Bladder: 0.92<br>Rectum: 0.84<br>HR CTV: 0.81 | 65% clinically acceptable, 33% minor edits, 2% major edits. |

| | | | | | | | |
|---|---|---|---|---|---|---|---|
| a low/middle-income country's experience[5]<br>Duprez D et al 2023 | | | | | | | |
| Comprehensive clinical evaluation of deep learning-based auto-segmentation for radiotherapy in patients with cervical cancer[6]<br>Chung SY et al 2023 | RTOG | EfficientNet-B0 U-Net | 16 | Targets: CTV1, CTV2, CTV3<br>OARs: Anorectum, Bladder, Spinal Cord, Cauda Equina, Right/Left Femoral Heads, Bowel Bag, Uterocervix, Liver, Left/Right Kidneys, Stomach, Duodenum | Train: 165; Val: 15 | OARs:<br>- High DSC (>0.90): Bowel Bag, Liver, Kidneys<br>- Moderate DSC (0.80–0.90): Bladder, Spinal Cord, Femoral Heads<br>- Low DSC: Stomach (0.67), Duodenum (0.73)<br>CTVs:<br>- CTV1/2/3: 0.75–0.80 | 66% accepted as-is, 34% required minor edits |
| Automatic segmentation for plan-of-the-day selection in CBCT-guided adaptive radiation therapy of cervical cancer[7]<br>Zhang C et al 2022 | N/A | nnU-Net (3D full resolution U-Net) | 4 | CTV, rectum, bladder, bowel bag | 23 patients, 272 CBCT images (Train-Test: 4-fold cross-validation) | CTV: 0.79, Bowel bag: 0.81, Rectum: 0.75, Bladder: 0.84 | 91.5% |
| RefineNet-based 2D and 3D automatic segmentations | RTOG | 2D RefineNet, 3D RefineNetPl | 6 | Target: CTV<br>OARs: Bladder, Small Intestine, Rectum, | 251 (Training), 31 (Validation), 31 (Testing) | CTV: 0.82<br>OARs:<br>- Bladder: 0.97<br>- Small Intestine: 0.95 | N/A |

| | | | | | | | |
|---|---|---|---|---|---|---|---|
| for clinical target volume and organs at risks for patients with cervical cancer in postoperative radiotherapy[8] Xiao C et al 2022 | | us3D, FCN, U-Net, CE-Net, UNet3D, ResUNet3D | | Right/Left Femoral Heads | | - Rectum: 0.91<br>- Femoral Heads: 0.98 | |
| A dual deep neural network for auto-delineation in cervical cancer radiotherapy with clinical validation[9] Nie S et al 2022 | N/A | SegNet (trained with multi-group data), U-Net | 6 | Target: CTV OARs: Bladder, Rectum, Bowel Bag, Right/Left Femoral Heads | Split: 121 (Training), 22 (Validation), 60 (Testing) | CTV: 0.85, Bladder: 0.93, Rectum: 0.84, Bowel Bag: 0.89, Right Femoral Head: 0.93, Left Femoral Head: 0.92 | 98% |
| Deep learning-based auto-segmentation of clinical target volumes for radiotherapy treatment of cervical cancer[10] Ma C et al 2022 | RTOG and JCOG | VB-Net | 3 | Clinical Target Volumes (CTV): pelvic lymph drainage area (dCTV1), parametrial area (dCTV2), postoperative CTV (pCTV1) | Training: 157, Validation: 20, Testing: 23 for dCTV1 and dCTV2, Training: 272, Validation: 30, Testing: 33 for pCTV1 | dCTV1: 0.88, dCTV2: 0.70, pCTV1: 0.86 | 63.5% required minor edits; 34.1% required significant edits. |
| Clinical evaluation of deep learning–based clinical target volume three-channel auto-segmentation algorithm for adaptive | RTOG and JCOG | TCAS (Three-Channel Adaptive Auto-Segmentation Network) | 1 | CTV1: Pelvic lymph drainage area (dCTV1/pCTV1) and parametrial area (dCTV2) | 92 (Training), 15 (Testing) | Method 1: 0.8155, Method 2: 0.8277, Method 3: 0.8914, Method 4: 0.8921 | N/A |

| | | | | | | | |
|---|---|---|---|---|---|---|---|
| radiotherapy in cervical cancer[11] Ma C ying et al 2022 | | | | | | | |
| Three-dimensional deep neural network for automatic delineation of cervical cancer in planning computed tomography images[12] Ding Y et al 2022 | Clinical experience | 3D V-net (compared with U-net) | 12 | Rectum, Sigmoid, Small bowel, Bladder, Pelvic bones, Colon, Spinal cord, Femoral Head (L/R), Kidney (L/R) | Training: 90 Validation: 10 Testing: 30 | Target:<br>- CTV: 0.85 (V-net) vs 0.83 (U-net)<br><br>OARs (V-net vs U-net):<br>- Rectum: 0.85 vs 0.84<br>- Sigmoid: 0.80 vs 0.72<br>- Small bowel: 0.79 vs 0.74<br>- Bladder: 0.94 vs 0.93<br>- Pelvic bones: 0.92 vs 0.92<br>- Colon: 0.82 vs 0.81<br>- Spinal cord: 0.73 vs 0.72<br>- Femoral Head_L: 0.82 vs 0.84<br>- Femoral Head_R: 0.81 vs 0.82<br>- Kidney_L: 0.92 vs 0.93<br>- Kidney_R: 0.92 vs 0.91 | N/A |
| A Feasibility Study of Deep Learning-Based Auto-Segmentation Directly Used in VMAT Planning Design and Optimization | N/A | 3D U-net | 9 | Target Not segmented (CTV evaluated for dose coverage) Bladder, Femoral Head (Left/Right), Kidney (Left/Right), Pelvic Bones, | Training: 105, Testing: 22 | OARs (3D U-net):<br>- Bladder: >0.94<br>- Femoral Head (L/R): >0.94<br>- Kidney (L/R): >0.94<br>- Pelvic Bones: >0.94<br>- Colon: 0.82<br>- Rectum: 0.83<br>- Spinal Cord: Part of range (0.82–0.96) | N/A |

| | | | | | | | |
|---|---|---|---|---|---|---|---|
| for Cervical Cancer[13] Chen A et al 2022. | | | | Spinal Cord, Colon, Rectum | | | |
| Improving predictive CTV segmentation on CT and CBCT for cervical cancer by diffeomorphic registration of a prior[14] Beekman C et al 2022 | N/A | 3D CNN with joint voxel-wise classification and registration of a prior | 1 | Target: Cervix-uterus CTV<br>OARs: Not specified | Training: 84 patients<br>(Testing split not explicitly stated) | CT: 0.87, CBCT: 0.80 | N/A |
| Automatic clinical target volume delineation for cervical cancer in CT images using deep learning[15] Shi J et al 2021 | Clinical experience | RA-CTVNet (3D UNet ) | 1 | Target: CTV<br>OARs: Not specified | Training: 64 patients<br>Validation: 10 patients<br>Testing: 10 patients<br>Total: 84 patients<br>(Additional 120 CBCT delineations included) | 3D Unet: 0.688 ± 0.107 3D ResUnet: 0.722 ± 0.088 3D SE ResUnet (CTVNet): 0.743 ± 0.079 A-CTVNet: 0.776 ± 0.071 RA-CTVNet: 0.792 ± 0.073 | N/A |
| Deep learning-based auto-segmentation of organs at risk in high-dose rate brachytherapy of cervical cancer[16] Mohammadi R et al 2021 | N/A | ResU-Net | 3 | Bladder, Rectum, Sigmoid | Training: 73 Validation: 10 Testing: 30 | Bladder: 95.7% ± 3.7% Rectum: 96.6% ± 1.5% Sigmoid: 92.2% ± 3.3% | N/A |
| Auto-segmentations by convolutional | N/A | 3D fully-convolutional CNN | 5 | Target: - Cervical: CTVNs OARs: | Training: 226 Testing: 40 | Cervical Cancer: - Femoral Heads (R: 0.94, L: 0.93) - Bladder: 0.84 | Cervical Cancer: - Femoral Heads: 80–90% Excellent |

| | | | | | | | |
|---|---|---|---|---|---|---|---|
| neural network in cervical and anorectal cancer with clinical structure sets as the ground truth[17]<br>Sartor H et al 2020 | | | | - Femoral Heads, Bladder, Bowel Bag | | - Bowel Bag: 0.88<br>- CTVNs: 0.82<br>Anorectal Cancer:<br>- Femoral Heads (R: 0.92, L: 0.91)<br>- Bladder: 0.94<br>- Bowel Bag: 0.83 | - Bladder: 40% Excellent, 30% Not Acceptable<br>- Bowel Bag: 40% Good, 40% Acceptable<br>- CTVNs: 40% Not Acceptable<br>Anorectal Cancer:<br>- Femoral Heads: 67–83% Excellent<br>- Bladder: 50% Excellent<br>- Bowel Bag: 27% Good, 33% Not Acceptable |
| Automatic contouring system for cervical cancer using convolutional neural networks[18]<br>Rhee DJ et al 2020 | RTOG | Hybrid CNN (Inception-ResNet-V2 for classification; 3D V-Net and 2D FCN-8s for segmentation) | 14 | - Targets (3): Primary CTV, Nodal CTV, PAN CTV<br>- OARs (11): Bladder, Rectum, Spinal Cord, Left/Right Femurs, Left/Right Kidneys, Pelvic Bone, Sacrum, L4/L5 Vertebral Bodies | Training: 2464 (2254 clinical + 210 KiTS19)<br><br>Testing: 170 (140 internal + 30 external) | Targets:<br>Primary CTV: 0.86<br>Nodal CTV: 0.81<br>PAN CTV: 0.76<br>OARs:<br>Bladder: 0.89<br>Rectum: 0.81<br>Spinal Cord: 0.90<br>Femur (L: 0.94, R: 0.93)<br>Kidney (L: 0.94, R: 0.95)<br>Pelvic Bone: 0.93<br>Sacrum: 0.91<br>L4 Vertebra: 0.91<br>L5 Vertebra: 0.90 | Clinical Acceptability:<br><br>OARs/Bony Structures: 97–100% acceptable.<br><br>CTVs: 70–87% acceptable (Nodal CTV had lowest acceptability at 70%) |
| Segmentation of organs-at-risk in cervical cancer CT images with a convolutional neural network[19]<br>Liu Z et al 2019 | N/A | Modified 2D U-Net | 7 | 7 OARs<br>- Bladder, Bone Marrow, Femoral Head (Left/Right), Rectum, Small Intestine, Spinal Cord | Training: 77<br>Testing: 14 | Bladder: 0.924<br>- Bone Marrow: 0.854<br>- Femoral Head (Left): 0.906<br>- Femoral Head (Right): 0.900<br>- Rectum: 0.791<br>- Small Intestine: 0.833<br>- Spinal Cord: 0.827 | 90.34% |

| | | | | | | | |
|---|---|---|---|---|---|---|---|
| Automatic segmentation of pelvic organs-at-risk using a fusion network model based on limited training samples[20]<br>Ju Z et al<br>2020 | Automated segmentation software | Dense V-Network | 5 | 5 OARs<br>- Bladder, Small Intestine, Rectum, Femoral Head, Spinal Cord | Training: 80<br>Testing: 20 | Bladder: >0.87<br>- Small Intestine: >0.87<br>- Rectum: >0.87<br>- Femoral Head: >0.87<br>- Spinal Cord: >0.87 | N/A |
